# Elevated FGF21 and triglycerides in *SLC25A13* carriers support the G3P-ChREBP Citrin Deficiency disease hypothesis: A cross-sectional biobank study

**DOI:** 10.64898/2025.12.10.25342016

**Authors:** Charles Brenner, Vinod Tiwari, Yi-Ming Chen, I-Chieh Chen

## Abstract

**Introduction:** People with Citrin Deficiency (CD), inactivated for both copies of *SLC25A13*, have lean metabolic dysfunction-associated steatotic liver disease (MASLD) and an aversion to sweets. The mouse model of CD, which inactivates *Slc25a13* and *Gpd2*, has been shown to elevate hepatic glycerol-3-phosphate (G3P) and activate carbohydrate response element binding protein (ChREBP) thereby increasing lipogenic transcription, which could account for MASLD, and circulation of FGF_21_, which could account for sweet aversion. The objective of this study was to determine if people with one or two copies of *SLC25A13* inactivated have higher circulating FGF_21_ and whether higher FGF_21_ is associated with higher circulating lipids in this population, consistent with a common driver of sweet aversion and lipogenesis.

**Methods:** Blood and urine from age- and sex-matched participants in the Taiwan biobank were selected and used to test the hypothesis that *SLC25A13* inactivation elevates circulating FGF_21_ and urinary sodium and to determine whether elevated FGF_21_ correlates with elevated circulating triglycerides in mutation carriers.

**Results:** Age-matched male and female mutation carriers have 2.2-fold and 2.8-fold higher circulating FGF_21_ than control individuals. CD patients and mutation carriers also had elevated urinary sodium and CD patients have greatly elevated circulating FGF_21_. Moreover, though higher FGF_21_ depresses circulating lipids in the general population, higher FGF21 was strongly correlated with higher circulating triglycerides in *SLC25A13* mutation carriers.

**Conclusion:** human data support insights gleaned from the mouse model of CD, that loss of *SLC25A13* drives both lipogenic transcription and circulation of FGF_21_.

**Key messages:**

* While mechanistic analysis of the mouse model of Citrin Deficiency suggested that carbohydrate response element binding protein is activated in the disease, thereby driving lipogenic gene transcription and expression of FGF21, no human data were available to validate this mechanism.
* The ccurrent study shows that Citrin Deficiency mutation carriers circulate high levels of FGF21 and, contrary to the general population, have a positive association between FGF21 circulation and circulating triglycerides. Two women with Citrin Deficiency also have very high levels of FGF21.
* This biobank study suggests that FGF21 and triglyceride synthesis may both be downstream of carbohydrate response element binding protein activation in people with the Citrin Deficiency gene mutations.

## Introduction

Citrin Deficiency (CD) is a rare autosomal disorder that disturbs hepatic protein, carbohydrate and lipid metabolism. Though the disease is pan-ethnic, the most common pathological alleles appeared in the Far East, where the allele frequency can be as high as 3%^1^. In the first month of life, CD is diagnosed by jaundice, hypercitrullinemia, hyperlactatemia and failure to thrive^2^. Diagnosis is confirmed as biallelic inactivation of *SLC25A13*, which encodes the glutamate-aspartate exchanging component of the malate aspartate shuttle (MAS)^3^. Highly expressed in hepatocytes, loss of *SLC25A13* leads to a deficiency in hepatocytosolic Asp required for argininosuccinate synthase 1, the urea cycle enzyme that links Asp to citrulline, thereby disturbing protein metabolism. Because the MAS is the principle means to move reducing equivalents from hepatocytosolic NADH to the mitochondrial matrix, CD patients are also largely carbohydrate intolerant. After dietary modification to a low protein, low carbohydrate diet rich in medium chain triglycerides (MCTs), failure to thrive usually resolves such that children with CD grow to adulthood^4^. Strikingly, they exhibit lean metabolic dysfunction-associated steatotic liver disease (MASLD)^5^ and an aversion to sweets^2^.

The leading mouse model of CD inactivates not only *Slc25a13* but *Gpd2*, a component of the glycerol-3-phosphate dehydrogenase shuttle (GDPS) that also moves cytosolic reducing equivalents to mitochondrial electron transfer. These mice recapitulate the major features of CD including lean MASLD, hypercitrullinemia and hyperlactatemia^6^. To better understand sweet aversion in CD, *Slc25a13* -/- *Gpd2* -/- mice were tested for preference of saccharine to water and behaved as wild-type with a large preference for the artificial sweetener. However, when offered sucrose, they developed an aversion to sucrose, ethanol and glycerol^7^.

FGF21 is a hepatokine and myokine expressed in a wide variety of seemingly paradoxical conditions of metabolic stress including both weight loss and obesity, mitochondrial dysfunction, fasting and refeeding, ketogenic diet and provision of simple carbohydrates (especially fructose), and ethanol^8,9^. Genetic and pharmacological data show that FGF21 acts through neuronal *KLB* receptors to cause aversion to sweets^10^ and alcohol^11^. We proposed that FGF21 is responsible for sweet aversion in people with CD^12^. In support of this, we discovered that mice with deletion of *Slc25a13* and *Gpd2* have elevated glycerol-3-phosphate (G3P), elevated hepatic expression of *Fgf21* mRNA, elevated circulation of FGF21 protein, and that G3P is a specific ligand of the carbohydrate-responsive N-terminal domain of the carbohydrate response element binding protein (ChREBP), whose activation drives *Fgf21* transcription^12^. The proposal that G3P activates ChREBP was notable in that, in the past, other glucose metabolites had been proposed to be the activating ligand and no previous model of FGF21 regulation proposed a mechanism to reconcile the apparent paradoxes of FGF21 induction^12^.

At the time of our mouse experiments, no data were available to determine if G3P leads to ChREBP activation, FGF21 circulation and *de novo* lipogenesis in human CD. However, we examined genome-wide association studies (GWAS) across databases that represent significant numbers of *SLC25A13* mutation carriers and discovered associations with low body weight, high urinary sodium and a preference for fatty fish, all of which could be indicators of elevated circulation of FGF21 in *SLC25A13* heterozygotes^12^. Here we used the Taiwan Biobank (TWB)^13^ to identify 25 male and 25 female *SLC25A13* mutation carriers and two female CD patients along with age- and sex-matched *SLC25A13* noncarriers to test the hypothesis that loss of *SLC25A13* in humans elevates circulation of FGF21. Further, based on the idea that FGF21 may be a circulating biomarker that reports on G3P-ChREBP signaling in *SLC25A13* mutation carriers and that G3P-ChREBP drives production of triglycerides (TGs) in people, we tested the hypothesis that FGF21 circulation is correlated with elevated TGs in *SLC25A13* heterozygotes. Here we show that loss of one copy of *SLC25A13* elevates FGF21 circulation and that the higher the FGF21 in *SLC25A13* mutation carriers, the higher the circulating TGs.

## Results

As shown in Fig. 1A and 1B, our data show that age-matched male and female *SLC25A13* mutation carriers have 2.2-fold and 2.8-fold higher circulating FGF21 than control members of the TWB with results significant at *P* = 0.056 and 0.012, respectively. To determine if we could confirm a GWAS association between *SLC25A13* mutation carriers and elevated urinary sodium^12^, we found that carriers have a 1.4-fold higher urinary sodium concentration. As shown in Fig. 1C, higher urinary sodium in carriers and CD patients with respect to controls was significant at *P* = 0.0052. These data suggest that the leanness, dietary preferences and elevated sodium of *SLC25A13* mutation carriers can be explained by elevated FGF21.

**Figure 1.**
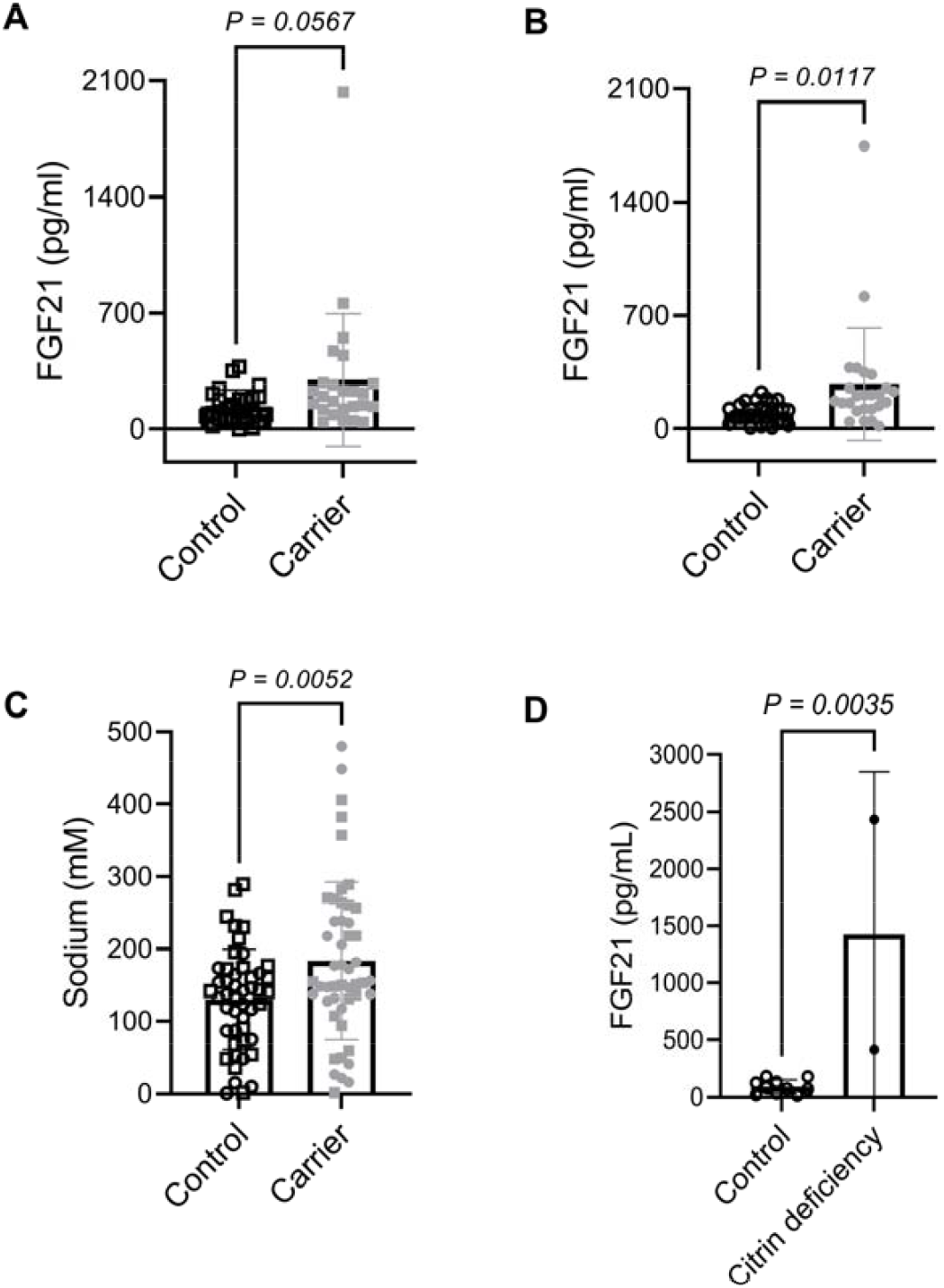
Loss of *SLC25A13* increases FGF21 circulation and urinary sodium. Age and ex-matched *SLC25A13* mutation carriers from the TWB were assayed for FGF21 circulation and urinary sodium. The data show that male (A) and female (B) mutation carriers have higher levels of FGF21 than control participants and that male and female mutation carriers have higher levels of urinary sodium (C) than control participants. Further, female CD patients have much higher levels of circulating FGF21 than control female age-matched participants (D). (A) Plasma FGF21 levels (N = 25 per group); (B) Plasma FGF21 levels (N = 27 controls, N = 25 carriers); (C) urinary sodium (N = 48 controls, N = 49 carriers); (D) plasma FGF21 (N = 10 controls, N = 2 CD). Symbols: □ male controls; ■ male carriers; ○ female controls; ●female carriers; ● female CD patients. *P* values are from Student’s t-test.

Across the 120,143 member TWB, the allele frequency for the 4 nucleotide *SLC25A13* deletion rs80338720 was 0.0052 and the expected number of homozygotes is three. We identified two CD patients in TWB (women of ages 57 and 61). As shown in Fig. 1D, we compared FGF21 levels in the two women with CD to FGF21 levels in the 10 women nearest their ages (*i*.*e*., 53 to 65). These data show that CD elevates FGF21 15-fold, *P =* 0.0035, consistent with the proposal that sweet aversion in CD is due to elevated FGF21^12^. Indeed, a clinically observable effect on sweet-liking versus sweet-disliking has been observed in healthy people with a less than 2-fold increase in fasting FGF21^14^.

Prior to our GWAS analysis^12^, there were no known health associations with *SLC25A13* carriers and our analysis indicated a signal for leanness. Nonetheless, we considered that loss of one copy of *SLC25A13* might increase accumulation of G3P sufficient to drive not only ChREBP-dependent transcription of FGF21 (Fig. 1A-B) but also the genes of TG synthesis. Though it is known that genetically proxied elevated FGF21 depresses circulating TGs in the general population^15^, we predicted that in *SLC25A13* carriers, FGF21 would serve as a biomarker of G3P-ChREBP signaling such that it will be positively correlated with TGs. TG levels were available for 7 of the 52 control TWB participants and for 45 heterozygotes. As shown in Fig. 2A, *SLC25A13* mutation carriers had 1.4-fold higher TG levels that did not meet statistical significance (*P* = 0.32) but which was directionally the same as the signal we observed by GWAS between *SLC25A13* carriers and elevated ApoB^12^. Moreover, as shown in Fig. 2B, consistent with the prediction that FGF21 expression in *SLC25A13* mutation carriers is a proxy of elevated G3P-ChREBP transcriptional activity, TGs show a strong positive association with FGF21 circulation in mutation carriers (Pearson correlation coefficient r = 0.58, *P =* 0.0001).

**Figure 2.**
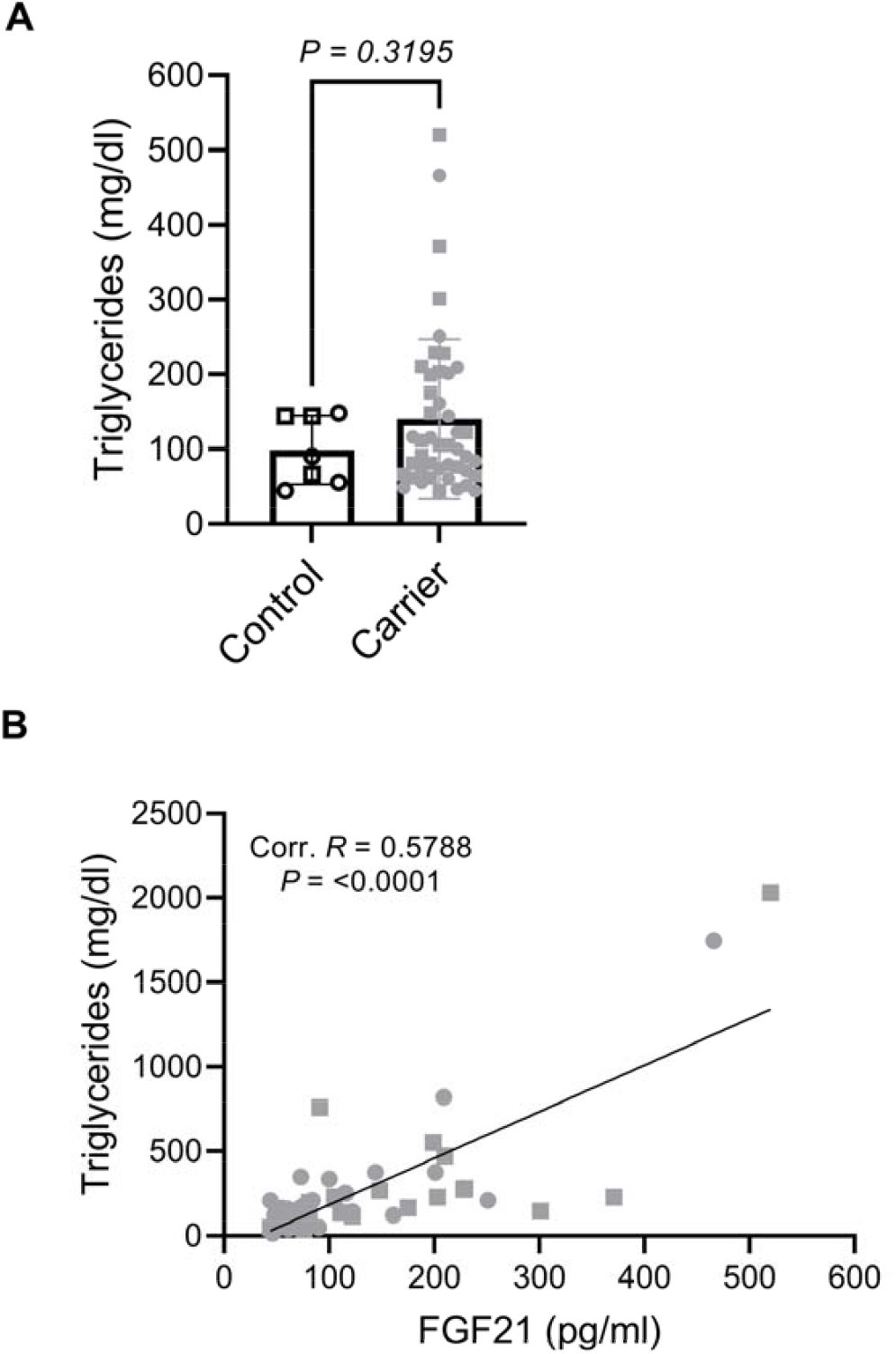
TGs are positively correlated with FGF21 circulation in *SLC25A13* mutation carriers. (A) TGs were compared between the N = 7 control participants and the N =45 *SLC25A13* mutation carriers for whom TG values were available in the TWB. These data show a nonsignificant TG increase in mutation carriers. (B) A plot of TGs as a function of FGF21 in mutation carriers shows a strong positive correlation (Pearson’s correlation R = 0.58) that is counter to the depression of TGs by genetically proxied FGF21 in the general population^15^ and consistent with elevation of FGF21 and lipogenic transcription by a potentiated ChREBP system in *SLC25A13* carriers.

According to our model, accumulation of G3P and consequent activation of ChREBP are at the root of two key features of CD. First, G3P-ChREBP drives a lipogenic gene expression program that turns on expression of pyruvate kinase, acetyl-coA lyase, acetyl-coA carboxylase, fatty acid synthase and elongation enzymes, and the Kennedy pathway enzymes that link newly synthesized long chain fatty acids (LCFA) to the G3P backbone to form TGs. Our mouse data further support the idea that G3P-ChREBP signaling depress expression and activity of carnitine palmitoyltransferase 1A (CPT1A), thereby accounting for the clinical benefit of MCTs rather than long chain TGs in nutrition for people with CD^12^. Second, activation of ChREBP by G3P would cause FGF21 to circulate. Our data establish that FGF21 circulation is increased by loss of one and both copies of *SLC25A13* in a manner that can account for sweet aversion. Further, consistent with the idea the FGF21 circulation reports on the degree of G3P-ChREBP transcription, our data show that FGF21 is strongly correlated to elevated TGs in *SLC25A13* mutation carriers.

## Discussion

The data are consistent with G3P-ChREBP driving both a lipogenic transcriptional program and a homeostatic FGF21 induction program that would tend to limit fructose and ethanol intake in order to block lipogenic energy inputs. Whereas Mendelian randomization shows that people with gain-of-function *FGF21* alleles have lower circulating TGs (and higher urinary sodium)^15^, loss of one copy of *SLC25A13* appears to sensitize to G3P-ChREBP activation and drive TG synthesis. In the context of biallelic *SLC25A13* inactivation and the resulting potentiated G3P-ChREBP signaling, we predict that the lipolytic effect of FGF21 on liver^16,17^ is insufficient to block MASLD. Further, as shown in Fig. 3, we propose that distinctive dual rise in both FGF21 and TGs in CD may potentially serve as a clinically useful biomarker with lower FGF21 reporting on disease modifiers such as potentially higher expression of the *SLC25A13-*paralogous *SLC25A12* gene^18^, which would increase Asp availability, lowering citrulline accumulation and lowering G3P to lower expression of FGF21 and the genes of *de novo* lipogenesis^12^. Higher FGF21 would potentially report on lack of *SLC25A12* expression and/or dietary components such as high carbohydrates, fructose and/or ethanol exposure that would elevate G3P, driving expression of FGF21 and the genes of *de novo* lipogenesis^12^. In support of this study, we and colleagues have shown that FGF21 is highly elevated in a 29 child cohort of pediatric CD and that FGF21 is positively correlated with levels of citrulline, threonine and TG^19^.

**Figure 3.**
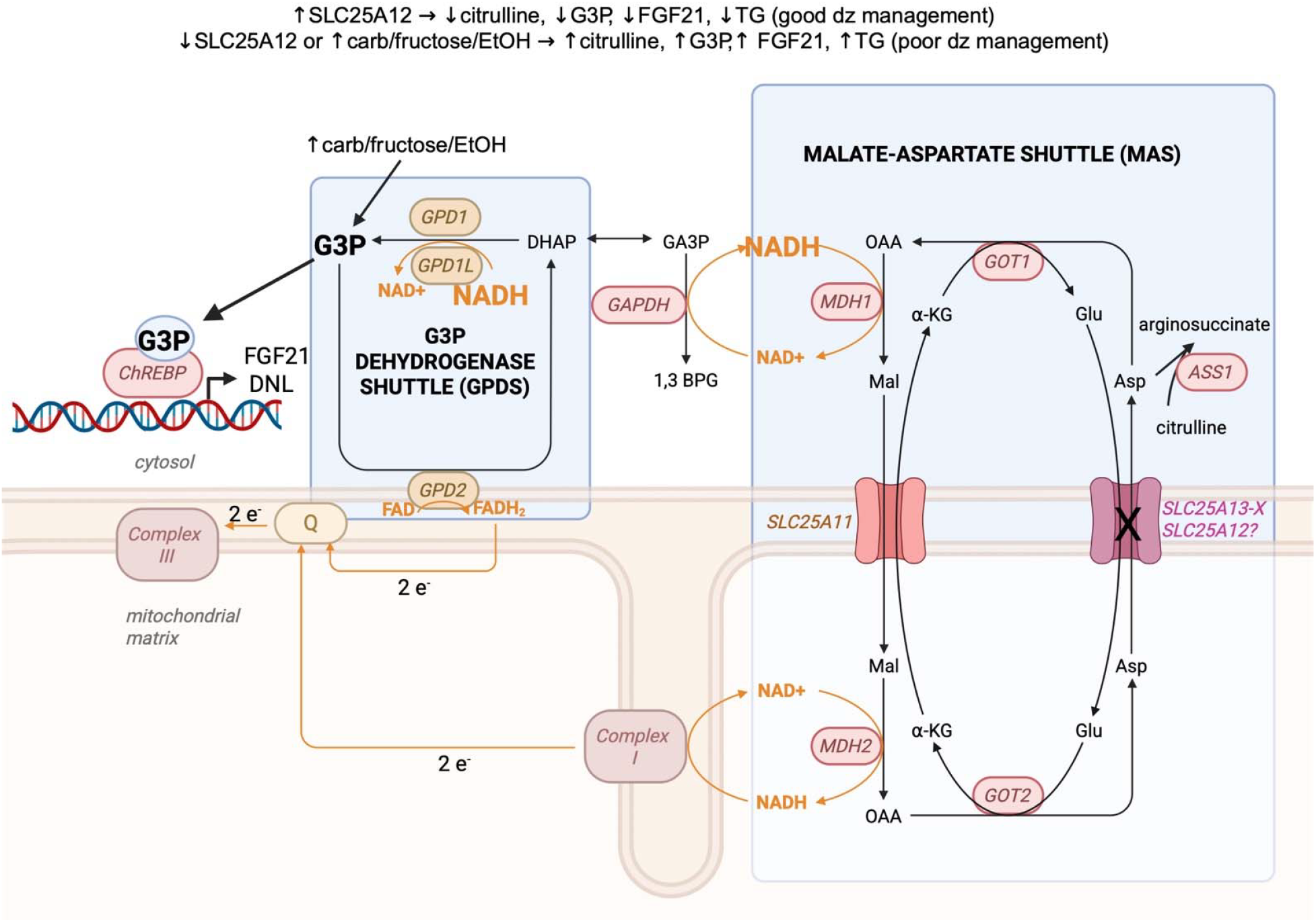
FGF21 as a potential reporter on CD disease management. The schematic shows flow of high energy electrons and Asp in hepatocytes as disrupted by loss of *SLC25A13*, encoding the major Glu-Asp exchanger in the mitochondrial inner membrane. Potential expression of *SLC25A12* would increase the supply of Asp to cytosol, preventing elevation of citrulline, NADH, G3P, FGF21 and lipogenesis. Lack of *SLC25A12* and dietary factors such as high carbohydrates, fructose and/or ethanol would tend to elevate G3P, thereby increasing ChREBP transcription of *FGF21* and the genes of *de novo* lipogenesis (DNL). Created in BioRender. Brenner, C. (2026) https://BioRender.com/uohluwl

This study has limitations. First, only two *SLC25A13* mutation carriers (both female) were identified, limiting statistical power and the ability to examine sex differences in biallelic CD. Second, the cohort was drawn exclusively from the Taiwan Biobank; replication in populations with greater genetic diversity is needed to confirm generalizability beyond East Asian ancestry. Nevertheless, the sex-independent effects of *SLC25A13* carrier status on FGF21 induction and correlation between FGF21 and TG in carriers provide support for the G3P-ChREBP hypothesis in humans. Now supported by human data validating the CD disease hypothesis, ongoing efforts are geared to query the role of G3P-ChREBP signaling in other conditions of elevated FGF21 expression and to test whether CPT1A activation will relieve lean MASLD in the mouse model of CD.

## Methods

Both sexes were included in this study, and sex was evaluated as a biological variable, though sex-specific results were not found. Biological samples and de-identified participant information were obtained by application to the TWB. The study protocol was reviewed and approved by the Institutional Review Board of Taichung Veterans General Hospital, Taiwan (IRB no. CE16270B). Participants provided written informed consent at enrollment, permitting use of their biospecimens and data for approved research. We scanned the TWB for entries positive for rs80338720, the 4 nucleotide deletion allele that is the most common pathogenic mutation in *SLC25A13*, and, in May 2025, requested biospecimens from the only two participants homozygous for this allele (both female), 25 women heterozygous for this allele, and 25 men heterozygous for this allele (52 cases), along with 27 age-matched women and 25 age-matched men (52 controls) within ±1 year of age. After study approval, urine and plasma samples were provided by the TWB. Plasma FGF21 levels were quantified in duplicate using the Quantikine® Human FGF-21 ELISA kit (R&D Systems, DF2100) according to the manufacturer’s protocol. Standards, controls, and plasma samples were assayed in duplicate. Urinary sodium concentrations were measured using the Sodium Assay Kit (Colorimetric; Abcam, ab211096). Urine samples were diluted 100–200x to fall within the linear range of the standard curve, and absorbance was read at 405 nm. For each group, the mean, standard deviation, median, and interquartile range of FGF21 (pg/mL) and sodium (mM) were calculated. Between-group comparisons were performed using Student’s t-test. Statistical analyses and data visualization were conducted with GraphPad Prism software. All data will be made available for analysis.

## Data Availability

Data will be made available in a public repository.

## Acknowledgements

The authors thank the Arthur Riggs Diabetes and Metabolism Research Institute for support, the Citrin Foundation for introductions and encouragement, and staff at TWB for their hard work in collecting and distributing data.

## Author Contributions

CB conceived of the study, provided reagents, interpreted data, and wrote the manuscript. Y-MC and ICC obtained IRB approval, performed assays and analyzed data. VT plotted data. All others approved the manuscript.

## Data

All deidentified data will be deposited in a structured public repository at the time of publication.

## Ethics approval

The study protocol was reviewed and approved by the Institutional Review Board of Taichung Veterans General Hospital, Taiwan (IRB no. CE16270B). Participants provided written informed consent at enrollment, permitting use of their biospecimens and data for approved research.

## References

1 Saheki, T., Moriyama, M., Funahashi, A. & Kuroda, E. AGC2 (Citrin) Deficiency-From Recognition of the Disease till Construction of Therapeutic Procedures. Biomolecules 10 (2020). 10.3390/biom10081100

2 Saheki, T. et al. Metabolic derangements in deficiency of citrin, a liver-type mitochondrial aspartate-glutamate carrier. Hepatol Res 33, 181–184 (2005). 10.1016/j.hepres.2005.09.031

3 Saheki, T. & Kobayashi, K. Mitochondrial aspartate glutamate carrier (citrin) deficiency as the cause of adult-onset type II citrullinemia (CTLN2) and idiopathic neonatal hepatitis (NICCD). J Hum Genet 47, 333–341 (2002). 10.1007/s100380200046

4 Saheki, T. & Song, Y. Z. in GeneReviews((R)) (eds M. P. Adam et al.) (1993).

5 Komatsu, M. et al. Citrin deficiency as a cause of chronic liver disorder mimicking non-alcoholic fatty liver disease. J Hepatol 49, 810–820 (2008). 10.1016/j.jhep.2008.05.016

6 Saheki, T. et al. Citrin/mitochondrial glycerol-3-phosphate dehydrogenase double knock-out mice recapitulate features of human citrin deficiency. J Biol Chem 282, 25041–25052 (2007). 10.1074/jbc.M702031200

7 Saheki, T. et al. Oral aversion to dietary sugar, ethanol and glycerol correlates with alterations in specific hepatic metabolites in a mouse model of human citrin deficiency. Mol Genet Metab 120, 306–316 (2017). 10.1016/j.ymgme.2017.02.004

8 Flippo, K. H. & Potthoff, M. J. Metabolic Messengers: FGF21. Nat Metab 3, 309–317 (2021). 10.1038/s42255-021-00354-2

9 Uebanso, T. et al. Paradoxical regulation of human FGF21 by both fasting and feeding signals: is FGF21 a nutritional adaptation factor? PLoS One 6, e22976 (2011). 10.1371/journal.pone.0022976

10 Jensen-Cody, S. O. et al. FGF21 Signals to Glutamatergic Neurons in the Ventromedial Hypothalamus to Suppress Carbohydrate Intake. Cell Metab 32, 273–286 e276 (2020). 10.1016/j.cmet.2020.06.008

11 Flippo, K. H. et al. FGF21 suppresses alcohol consumption through an amygdalo-striatal circuit. Cell Metab 34, 317–328 e316 (2022). 10.1016/j.cmet.2021.12.024

12 Tiwari, V. et al. Glycerol-3-phosphate activates ChREBP, FGF21 transcription and lipogenesis in citrin deficiency. Nature Metabolism 7, 2284–2299 (2025).

13 Feng, Y. A. et al. Taiwan Biobank: A rich biomedical research database of the Taiwanese population. Cell Genom 2, 100197 (2022). 10.1016/j.xgen.2022.100197

14 Soberg, S. et al. FGF21 Is a Sugar-Induced Hormone Associated with Sweet Intake and Preference in Humans. Cell Metab 25, 1045–1053 e1046 (2017). 10.1016/j.cmet.2017.04.009

15 Giontella, A. et al. Renoprotective effects of genetically proxied fibroblast growth factor 21: Mendelian randomization, proteome-wide and metabolome-wide association study. Metabolism 145, 155616 (2023). 10.1016/j.metabol.2023.155616

16 Inagaki, T. et al. Endocrine regulation of the fasting response by PPARalpha-mediated induction of fibroblast growth factor 21. Cell Metab 5, 415–425 (2007). 10.1016/j.cmet.2007.05.003

17 Badman, M. K. et al. Hepatic fibroblast growth factor 21 is regulated by PPARalpha and is a key mediator of hepatic lipid metabolism in ketotic states. Cell Metab 5, 426–437 (2007). 10.1016/j.cmet.2007.05.002

18 Mention, K. et al. Citrin deficiency: Does the reactivation of liver aralar-1 come into play and promote HCC development? Biochimie 190, 20–23 (2021). 10.1016/j.biochi.2021.06.018

19 Liang, J. et al. FGF21 Elevation is a Biomarker of Citrin Deficiency and its Metabolic Dysregulation in Pediatric Patients. in review (2026).

